## Supplemental material for "You are where you eat: Effect of mobile food environments on fast food visits"

#### Supplementary Notes

**1 Data** **4**

**2 Representativity** **4**

**3 Classification of Food Outlets** **5**

**4 Definition of context and food visit** **6**

**5 Detecting change of context** **7**

**6 Estimation of the effect of changing contexts** **8**

**7 Models** **9**

**8 Topic analysis of census tracts** **11**

**9 Effect of interventions on other health groups** **12**

**10 Robustness checks** **13**

**11 Interventions** **15**

**12 Libraries used** **15**

### List of Figures

S1 A: Correlation between the smartphone population detection in our mobility data and the official census population by census tract. B: Coefficients of the logistic regression coefficient without (raw) and with (weighted) weights in the regression proportional to $1/w_\Omega$. C: Effect of the different interventions re-weighted by $1/\Omega$ to account for the different ratio of users to population in each census tract $\Omega$. . . . . . . . . . . . . . 6
S2 Comparison between the number of FFO by chain in different cities obtained from our Foursquare dataset and Infogroup's US Historical Business Data for 2016 [14]. . . . 7
S3 Method to detect the change of context before lunch. We construct a binary time series for each of the census tracts for those users that have more than 30% of their contexts in two of them. We use a statistical test to detect statistically significant changes in the mean of those time series to know if and where changepoints happened. For the user in panel A, a significant change in the means (in red) is detected in December. However, for the user in panel B, no significant change is detected, and the user changes often between contexts. Statistically significant changes were detected using the `changepoint` library with a manual method and penalty of $n^{1/10}$ (see details of those methods in [15]). . . . . . . . . . . . . . . . . . . . . . . . . . . . . . . . . . 8
S4 Left: Effect of the mobile food context $\phi$ in choosing an FFO using a logistic regression and hot-encoding different values of $\phi$. Error bars are standard errors of the coefficients. Right: Effect of the mobile food context in choosing an FFO restricted to the food outings of different socio-demographic groups. Groups are selected as quantiles of each socio-demographic variable. . . . . . . . . . . . . . . . . . . . . . 11
S5 Importance of the FFO context variable $\phi(\mathbf{c}_{it})$ for models built with actions at different hours of the data. Variable importance is given by the absolute value of the $t$-statistic of the variable in the regression model (2) by the hour. . . . . . . . . . . . . . . . . 12
S6 Variation of different metrics to estimate the optimal number of topics in the different census tracts. Metrics were obtained using the `ldatuning` package [18]. As we can see, several metrics show a minimum/maximum around $k = 20$. . . . . . . . . . . . 13
S7 Composition of the different $k = 20$ POI topics found in our census tracts. We have given names to each of them for easy recognition. Bars represent the probability of finding each POI category in each of the topics. Note: no mobility data or visits were used in this analysis, only the spatial distribution of POIs from the Foursquare API. . 21
S8 Average (and standard error of the mean) frequency of each topic across all census tracts in our dataset. . . . . . . . . . . . . . . . . . . . . . . . . . . . . . . . . . . 22
S9 Effect of the intervention strategies on the different health outcomes groups. Each panel shows the total increase in the number of non-FFO visits by the changed actions of users belonging to different health outcomes groups (shades). We classify each individual into different groups of health outcomes depending on which quantile their census tract is with respect to that outcome (darker shading for higher quantiles of each health outcome group). Health outcomes by census tract are obtained from the PLACES Local data for Better Health dataset from the CDC [5]. . . . . . . . . . . 23

S10 Effect of the mobile phone context in choosing a FFO when going to a food place at lunchtime for different definitions of FFO for different attributions of the stays to the POI and the choice of the stay for the context before lunch. Values show the coefficient $\beta$ of the logistic regression (log-odds) in Eq. [2] and bars indicate its standard deviation. The result in the main paper corresponds to 200m, within a 1km radius and last stay. . . . . . . . . . . . . . . . . . . . . . . . . . . . . . . . . . 24
S11 Left: Effect of the mobile phone context in choosing a FFO when going to a food place at lunchtime for different definitions of FFO. The analysis is only made for the Los Angeles metropolitan area since the manual definition in [13] is only available there. Values show the coefficient of the logistic regression (log-odds) in Eq. [2] and bars indicate its standard deviation. Right: Number of FO and FFO in the different definitions of fast food outlets. . . . . . . . . . . . . . . . . . . . . . . . . . . . . 24

S4 Results for the Ordinary Least Squares regression of mobile ($\phi_i^m$), home ($\phi_i^h$) average exposure to FFO environments, and the fraction of FFO visits ($\mu_i$) by individual, as a function of different socio-demographic traits of the census tract where individuals live. 19
S5 Logistic regression results for the model in Equation [2] for different groups of users and food outings. The last column corresponds to the model in Equation [3] for the DMV visits. Numbers in parenthesis correspond to standard errors of the coefficients. We also report the Squared Correlation and Pseudo $R^2$ results for the logistic regression. . . . . . . . . . . . . . . . . . . . . . . . . . . . . . . . . . . . . . . . 20

### 1 Data

The mobility data were obtained from Cuebiq, a location intelligence, and measurement company. The dataset consists of anonymized records of GPS locations from users who opted-in to share the data anonymously in the US metropolitan areas over a period of 6 months, from October 2016 to March 2017. Data was shared in 2017 under a strict contract with Cuebiq through their Data for Good program, where they provide access to de-identified and privacy-enhanced mobility data for academic research and humanitarian initiatives only. All researchers were contractually obligated to not share data further or to attempt to de-identify data. Mobility data is derived from users who opted in to share their data anonymously through a General Data Protection Regulation (GDPR) and California Consumer Privacy Act (CCPA) compliant framework.

From the data, we extracted the "stays", as the places $\mathbf{x}_{i,t}$ where anonymous user $i$ stayed (stopped) at time $t$ for at least 5 minutes using the algorithm proposed by Hariharan and Toyama [11]. We also calculated the duration of the stay, $\tau(\mathbf{x}_{i,t})$. To discard stays related to working places, we only considered stays of less than 2 hours of duration [$\tau(\mathbf{x}_{i,t}) < 2$ hours]. Some of the stays happen within places (Points of Interest). We use a dataset of 1.2 Million Points of Interest in US metropolitan areas collected using the Foursquare API in 2017. We use the Foursquare venue categorization of the places to detect the type of place visited [9]. Finally, we estimate the home Census Block Group of the anonymous users as that in which they are more likely located during nighttime. This results in a dataset of the places people stayed, including the points of interest that anonymous users visited and the most likely census block group of where the device owner lives. We also discard stays happening closer than 50 meters from users' home location.

We only considered mobility data that happened within 11 metropolitan areas defined as the Core-based Statistical Areas (CBSA) [1]. We considered CBSAs instead of other geographical units since they are areas that are socioeconomically related to an urban center. This provides a self-contained metropolitan area in which people move for work, leisure, or other activities. Note that most of the CBSAs we consider span several states.

Table S1 shows a summary of the data used in the analysis. More details about how the data was constructed, its representativity, and comparison with other datasets can be found in [17, 13] and Section 2. In particular, as found in [17] our sample of users and their behavior is highly representative of the people living in those metropolitan areas, including their socio-demographic profile and the visitation patterns to different amenities. Also in [13] we found that neighborhood-level features representing visits to fast food outlets (FFO) were significantly associated with self-reported fast-food intake, significantly associated with obesity and diabetes, and were a better predictor of these diseases than self-reported fast food intake.

### 2 Representativity

Our location data comes from smartphones in large urban areas. Although a large proportion of the U.S. population owns a smartphone in urban areas, we might question whether our sample of 1.8 million devices is representative of the population and different socio-demographic groups in that area. Figure S1 shows the comparison between the population detected in our data and the census data. As we can see, the correlation is high ($\rho = 0.61 \pm 0.01$) by census tract, showing that we get a good representation of the population. Despite that, similar to reference [17] we address the representativity of the data using a weighting mechanism (post-stratification) based on $w_\Omega$, the ratio

**Table S1:** Summary of the mobility data and visits in each metro area in thousands (k) or millions (M).

| Metro Area | Number of users | Number of total visits | Number of food visits (% of total visits) | Number of fast food visits (% of total food visits) |
|---|---|---|---|---|
| Boston | 72k | 7.49M | 2.08M (27.79%) | 0.3M (14.45%) |
| Chicago | 300k | 39.18M | 9.46M (24.14%) | 1.85M (19.59%) |
| Dallas | 245k | 33.43M | 8.13M (24.32%) | 1.89M (23.27%) |
| Detroit | 127k | 15.91M | 3.91M (24.58%) | 0.71M (18.16%) |
| Los Angeles | 243k | 38.37M | 9.92M (25.85%) | 1.72M (17.38%) |
| Miami | 187k | 26.63M | 6.29M (23.63%) | 0.93M (14.7%) |
| New York | 248k | 35.39M | 9.3M (26.26%) | 1.19M (12.85%) |
| Philadelphia | 123k | 13.68M | 3.7M (27.08%) | 0.51M (13.84%) |
| San Francisco | 86k | 9.6M | 2.65M (27.64%) | 0.37M (13.98%) |
| Seattle | 73k | 8.58M | 2.33M (27.16%) | 0.43M (18.44%) |
| Washington | 153k | 16.77M | 4.26M (25.41%) | 0.9M (21.01%) |
| Total | 1862k | 245.05M | 62.04M (25.32%) | 10.81M (17.42%) |

of smartphone users to the true population in census tract $\Omega$ [21]. We tested that our main results do not depend on this post-stratification technique. In particular, as we can see in figure S1b if we weight the data in the logistic regression model (2) by the inverse of the ratio, we get very similar results to the raw data. Still, the impact of the environment is of the same magnitude as the main results presented in the paper. Similar results are obtained when we re-weight the effect of different interventions by the inverse of the ratio of smartphone users to the true population. In Figure S1c we compare the effect on different interventions on our sample of users, compared with the weighted version

$$\Delta^{\mathrm{FFO}}(\Omega) \simeq \sum_{\mathbf{c}_{it}\in\Omega} \frac{\beta}{w_\Omega} \frac{e^{X_{it}}}{(1+e^{X_{it}})^2} \frac{\delta\phi}{\delta \mathrm{I}}.$$

As we can see, the relative results between the different interventions are similar between the weighted and non-weighted (raw) data. But obviously, the numbers are scaled to population levels.

#### 3 Classification of Food Outlets

Food visits were defined as visits to a location where food might be sold (including restaurants, food retailers, and other locations). We extend the idea in [7] and identify Food Outlets (FO) using a combination of Foursquare's existing categorization taxonomy [9]. Each place in the Foursquare dataset belongs to a parent category (e.g., "Food" or "Arts & Entertainment"). In particular, FO are defined as those in the "Food" parent category, but we also include other places that serve or sell food which happen in other parent categories, probably due to misclassifications or FO within multipurpose facilities (e.g., music venue that serves food). A detailed description of those combinations appears in Table S2. For privacy reasons, we have excluded FO located at schools, drug stores or medical facilities.

To identify Fast Food Outlets (FFO) we use the name of the venue and match it to the list of Fast Food restaurant chains in the US from the Wikipedia `https://en.wikipedia.org/wiki/List_of_fast_food_restaurant_chains#United_States`. We augmented that list to get common names like "KFC" instead of the official "Kentucky Fried Chicken". And to account for

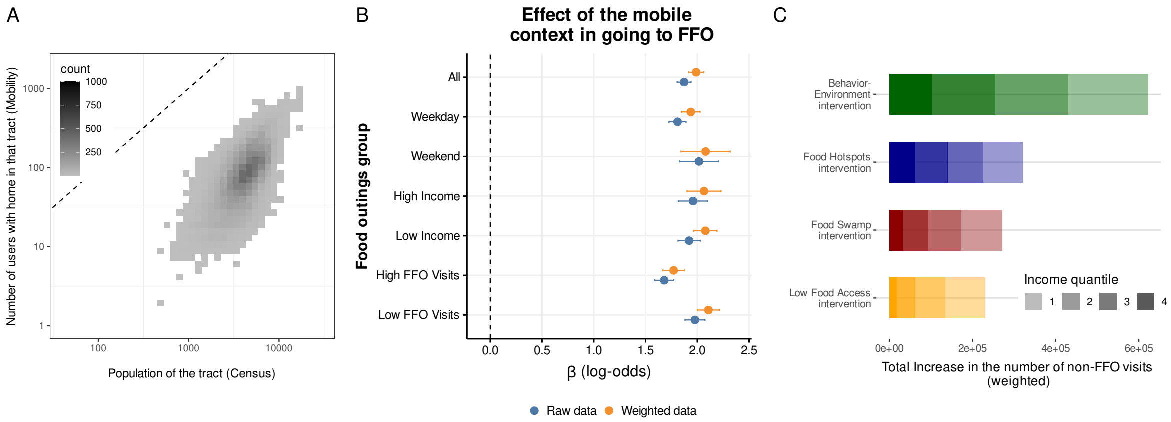

**Figure S1:** A: Correlation between the smartphone population detection in our mobility data and the official census population by census tract. B: Coefficients of the logistic regression coefficient without (raw) and with (weighted) weights in the regression proportional to $1/w_\Omega$. C: Effect of the different interventions re-weighted by $1/\Omega$ to account for the different ratio of users to population in each census tract $\Omega$.

potential different spellings of the name (e.g. "Carl's Jr" instead of "Carl's Jr. / Green Burrito") we used approximate string matching between the name of the venue and the list in the Wikipedia using the Jaro-Winkler distance [6] of the `stringdist` library [24]. To test how good is this classification of FFO, we compare it with the manual annotation of FFO for the LA metro area in [13], which used a combination of Foursquare's existing FFO categorization taxonomy and a bottom-up search of known chain FF outlet names validated in previous research [8] and string matching techniques. We found that 94% of the places annotated as FFO in [13] were included in our automatic classification. However, our classification contains more FFO (6964) than those in [13] (4151) in LA, because some major chains like "Chipotle" or "Starbucks" where not considered FFO in [13]. In Section 10 we check extensively that our results do not depend on the definition and set of FFO considered.

Finally, our Foursquare dataset is an accurate representation of the fast-food outlets in each city. As we can see in Figure S2, the number of FFO detected in each city by fast-food chains has a strong correlation with other (curated) datasets of business, like the Infogroup's US Historical Business Data for 2016 [14]. The small variation between them can be attributed to the different way an outlet is defined in both datasets. Also, in our dataset, we can detect food outlets within businesses (e.g. coffee shops in large stores) which do not appear in Infogroup's dataset.

### 4 Definition of context and food visit

To understand the role of individual preferences and the context where food decisions are made, we restrict the analysis to visits to FO during lunch hours (from noon to 13h30 local time) because they are more frequent, more likely to involve a visit to a venue for food, but most importantly, because they are more likely to be affected by the local context where individuals were previously. As we can see in Figure 2 in the main text, visits to FO and FFO are more frequent around lunchtime. For those food decisions $y_{it}$ at time $t$, we take the last place in the morning (until 11:30) where user were

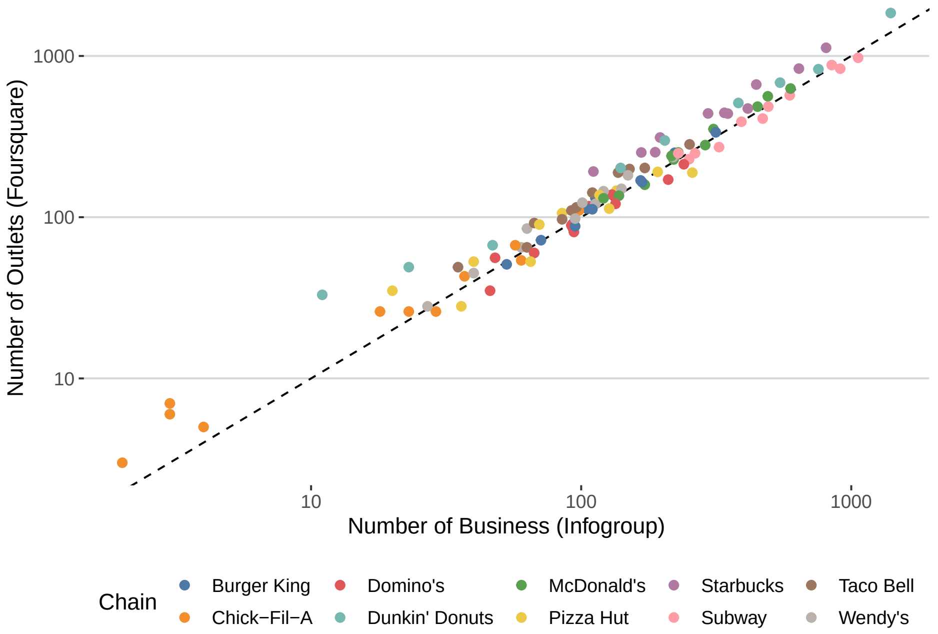

**Figure S2:** Comparison between the number of FFO by chain in different cities obtained from our Foursquare dataset and Infogroup's US Historical Business Data for 2016 [14].

observed (context $\mathbf{c}_{it}$), and characterize the context where that decision is made by $\phi(\mathbf{c}_{it})$. To make sure that there was a possible decision between FFO and non-FFO options, we only keep observations where the context $\mathbf{c}_{it}$ contains both type of places. After those restrictions, we got 2.7 million visits to FO during lunchtime by 755k users.

In our model, we have considered decisions $y_{it}$ which are done away from the pre-lunch context. For our lunchtime dataset, the distance from home to the FO visit has a median of 9.78km, while the median from the context is only 1.60km. Despite that, we get a number of actions $y_{it}$ that happen far away from the context. Note that if we restrict too much the set of actions to get only those that happen close to the context, we might end up with large endogeneity in our models. This is due to the fact that if the action happens very close to the context, indeed, the variable $\phi(\mathbf{c}_{it})$ will be larger from those actions $y_{it} = 1$ just because it includes the FFO where it happens. In any case, we have also run our models restricting the actions to only those that happen below a given distance $d^{max}$ from the pre-lunch context. As expected (see Table S3), due to the endogeneity, the log-odds $\beta$ are larger for smaller distances, but the models have similar predicting power, and the effect of the environment is always in the same direction.

#### 5 Detecting change of context

To detect those users that change their context before lunch we used the following method: we take the sequence of the census tracts where $c_{it}$ is. We only take those users that have at least two different census tract contexts with more than 30% of their total visited contexts during that time period, and we consider only users that appear at least 5 times at each of those two tracts. With only those two

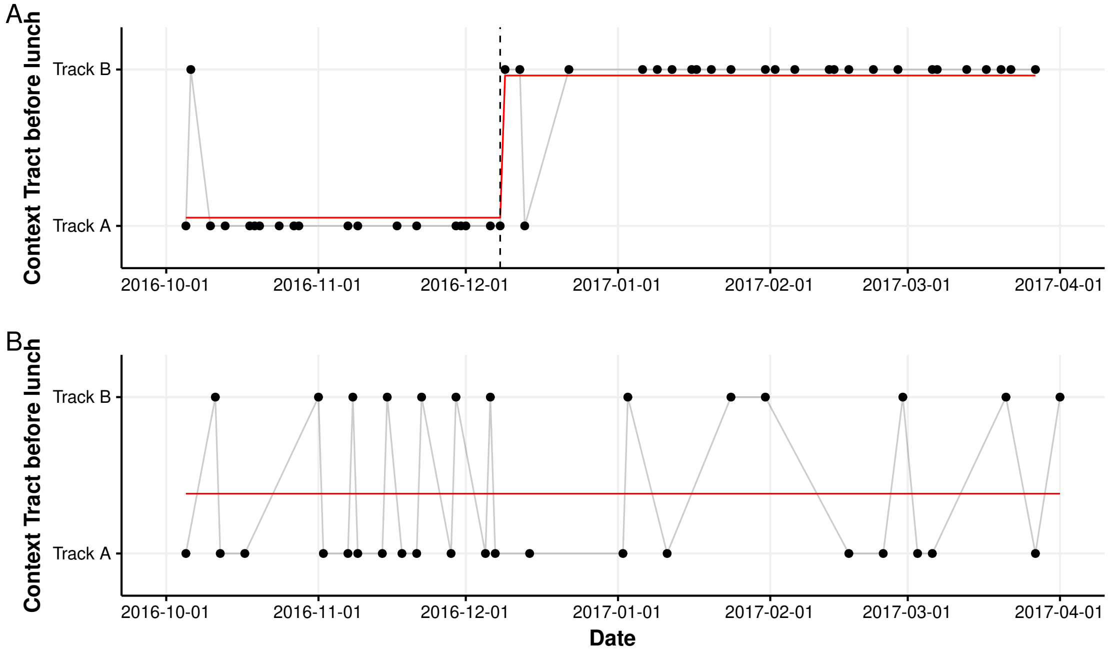

**Figure S3:** Method to detect the change of context before lunch. We construct a binary time series for each of the census tracts for those users that have more than 30% of their contexts in two of them. We use a statistical test to detect statistically significant changes in the mean of those time series to know if and where changepoints happened. For the user in panel A, a significant change in the means (in red) is detected in December. However, for the user in panel B, no significant change is detected, and the user changes often between contexts. Statistically significant changes were detected using the `changepoint` library with a manual method and penalty of $n^{1/10}$ (see details of those methods in [15]).

census tract contexts, we construct the binary time series between them, see Figure S3. We apply a method to detect if and where a changepoint occurred in that time series based on detecting statistical changes in the mean using the [15]. That method was implemented using the library `changepoint` in `R`, and we use a manual penalty using $n^\gamma$ for the number of changepoints $n$. We manually annotated some cases and found that $\gamma = 0.1$ yielded the best accuracy in detecting changes of contexts. If the method detects a changepoint, the properties of the mobile food environment before and after it are averaged over all $\mathbf{c}_{it}$ in each of the census tracts' contexts. Using this method, we detected 7913 users in our dataset that changed contexts during our observation period.

### 6 Estimation of the effect of changing contexts

To provide a statistically robust estimation of the impact of those individuals that changed their context before lunch, we design a methodology based on the Bayesian Structural Time Series (BSTS) approach. We define four groups of users depending on whether their contexts before/after has Low ($\phi < 0.13$) or High ($\phi > 0.13$) (average) ratio of fast food options. Around 34% of users changed from Low $\rightarrow$ Low and another 34% from High $\rightarrow$ High, while 16% changed from Low $\rightarrow$ High and another 16% from High $\rightarrow$ Low. To study the effect of changing from Low $\rightarrow$ High, for example, we consider the time series $\mu_t^{\text{Low}\rightarrow\text{High}}$ of the fraction of FFO visits at lunch by day $t$, where $t = 0$ is when they changed their context. As a counterfactual, we consider those users that changed their context also, but changed to a similar number of FFO options contexts, $\mu_t^{\text{Low}\rightarrow\text{Low}}$. Note that we did not use as counterfactual those users that stayed in the same geographical context, but only those that changed their geographical context. This was done to reduce the possibility of some endogeneity between changing contexts and the food environment in the previous context. Similarly to study the effect of changing from High $\rightarrow$ Low contexts we use $\mu_t^{\text{High}\rightarrow\text{Low}}$ taking $\mu_t^{\text{High}\rightarrow\text{High}}$ as counterfactual. BSTS models were implemented using the `CausalImpact` packages in `R` [4]. Given a response and a counterfactual time series, BSTS predicts how the response would have evolved after the change to a different context if the change never happened. In the modeling, we used the default values of the `CausalImpact` library, including no seasonality, since our time series are centered 50 days around where they change their context.

### 7 Models

#### 7.1 Demographic models for exposure to home and mobile food environments and FFO visits

To investigate the socio-demographic differences in home and mobile exposure to fast food, and in the fraction of FFO visit decisions, we have used simple linear regression models like

$$\phi_i^m, \phi_i^h, \mu_i \sim \sum_l \beta_l d_{l,i} + \text{MSA}_i \tag{1}$$

where $d_{l,i}$ are the different socio-demographic variables associated with the home Census Block Group (CBG) where user $i$ lives, and $\text{MSA}_i$ is a fixed factor by urban area. The socio-demographic variables considered come from the American Community Survey (5 years) 2017 edition, [23] and they correspond to (by CBG):

- Proportion of Black population.
- Proportion of employed (civilian) population.
- Proportion of people with college education or beyond.
- Proportion of people working in low-skill jobs, namely in the Agriculture, Construction, Manufacturing, Wholesale, Retail, and Transportation sectors.
- Proportion of people commuting longer than 45 minutes.
- Proportion of people that use public transportation for that commuting.
- Median income of each household.

Other variables (like the proportion of White people) were discarded as they are highly correlated with combinations of those above. Table S4 shows the results of the linear regression for each variable. As we can see, despite many of the coefficients being significant, only the model for $\phi^m$ has some explanatory power with $R^2 = 0.21$. Interestingly, the fraction of FFO visits and exposure at home models do have a very small explanatory variable.

##### 7.2 Model for visits to FO during Lunchtime

To test for the effect of FFO environments, we have run a number of statistical models. For the main results of the paper, we establish the effect of FFO environments around the contexts to go to a FFO place using a logistic regression for the probability that the user $i$ went to a FFO at time $t$, i.e. $y_{it} = 1$.

$$\Pr(y_{it} = 1) = \text{logit}^{-1}[\beta_0 + \alpha_i + \delta_t + \beta\phi(\mathbf{c}_{it})] \tag{2}$$

where $\text{logit}^{-1}(x) = e^x/(1 + e^x)$, and $\phi(\mathbf{c}_{it})$ is the fraction of FFO options around the context before lunch. We control individual preferences and daily patterns by introducing a fixed effect by user ($\alpha_i$) and day ($\delta_t$). Regression was only performed for those individuals or days that have at least one FFO and non-FFO visit. Results for this model are presented in Figure 2 of the main paper and Table S5. We also cluster errors by day to account for potential heterogeneity. However, our results are largely independent of that clusterizationS5.

To observe potential non-linearities in the effect of FFO, we used a logistic regression where we one-hot encoded the rounded version of $\phi(\mathbf{c}_{it})$. This allows us to observe whether the effect they have is linear, starts big and then plateaus, or it has another shape. The coefficient for each value of $\phi$ can be seen in figure S4. The effect is always increasing, and it is almost linear, as we hypothesize in Equation [2].

##### 7.3 Model for visits to FO by socio-demographic traits

We have also tested whether the effect of the context could be accounted for the different contexts that individuals visit depending on their socio-demographic traits. As we can see in Figure 2 in the main paper, people that rely more on public transportation and long commuting, with low-skill jobs, and from predominantly Black neighborhoods seem to be exposed to more FFO. However, as we see in Figure 2 and Figure 3 in the main paper, the FFO context and its effect seem to be largely independent of income. Rather than introducing complicated interaction terms, to test other demographic variables in the effect of the context, we have rerun the logistic regressions only for the visits to FO of particular socio-demographic groups, specifically by quantile of the different socio-demographic variables. As we can see in Figure S4, although there is some variability, our estimations of $\beta$ are very similar across different socio-demographic groups.

##### 7.4 Model for the visits to FO after DMV

In the case of visits to DMV, we typically have only one observation and day per user. Thus we have removed the daily and individual fixed factors and we have used a simpler logistic regression model:

$$\Pr(y_{it}^{\text{DMV}} = 1) = \text{logit}^{-1}[\hat{\alpha}_i + \beta\phi(\mathbf{c}_{it})] \tag{3}$$

where $\hat{\alpha}_i$ is the observed fraction of visits to FFO for each individual $i$.

##### 7.5 Model for the visits to FO during the whole day

Finally, we have extended model (2) to the rest of the day. To this end, we consider each stay in our dataset as a context $\mathbf{c}_{it}$, and we evaluate if there is a food visit $y_{it}$ in the next two hours after that stay. Then we fit a similar model as that in Eq. (2) only for those stays that include a visit to a food outlet in the next two hours. This allows us to investigate also at what time of the day is more important the

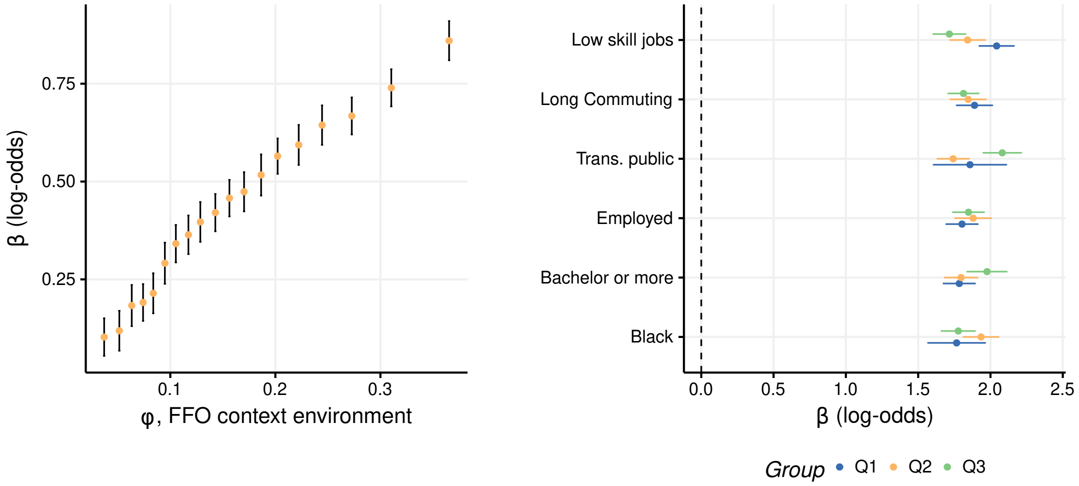

**Figure S4:** Left: Effect of the mobile food context $\phi$ in choosing an FFO using a logistic regression and hot-encoding different values of $\phi$. Error bars are standard errors of the coefficients. Right: Effect of the mobile food context in choosing an FFO restricted to the food outings of different socio-demographic groups. Groups are selected as quantiles of each socio-demographic variable.

context in the type of food visit. As we can see in figure S5, the importance of the context variable $\phi(\mathbf{c}_{it})$ measured by the t-statistic in the regression model (2) is larger around noon. Thus, visits around noon are more likely to be affected by the local context where individuals were previously.

#### 8 Topic analysis of census tracts

To identify the type of areas where the most efficient interventions happen, we have used topic modeling to identify the different patterns behind the distribution of POIs by area. We note that mobility data or visits were not used in this analysis, only the spatial distribution of POIs from the Foursquare API. Similar to LDA applied to text classification, we describe every census tract by the number of POIs in each category (terms), and we have applied Latent Dirichlet allocation (LDA) to get a small number of groups of POIs (topics) to describe each census tract (document) [3]. For our analysis, we have only considered those categories that appear at least in 20 or more census tracts, so we ended up with a document-term matrix of 18898 census tracts with 614 categories of POIs.

The result of the LDA applied to that document-term matrix are two matrices: one containing the weights of the different categories by topics and the other containing the probabilities of finding a specific topic within each census tract. To find the number of topics, we have used different metrics found in the `ldatuning` package [18]. Results for this tuning are found in figure S6 where we can see that we get (almost) global minima and maxima for different metrics around $k = 20$.

The composition of the different topics can be found in figure S7. As we can see, the topics are easily recognizable, and we have given names to each of them for easy recognition. They go from groups of POIs related to airports, to offices, industrial / factory areas, shopping centers, malls, health areas, education, and recreational areas, to more local ones related to different types of neighborhood groups of POIs.

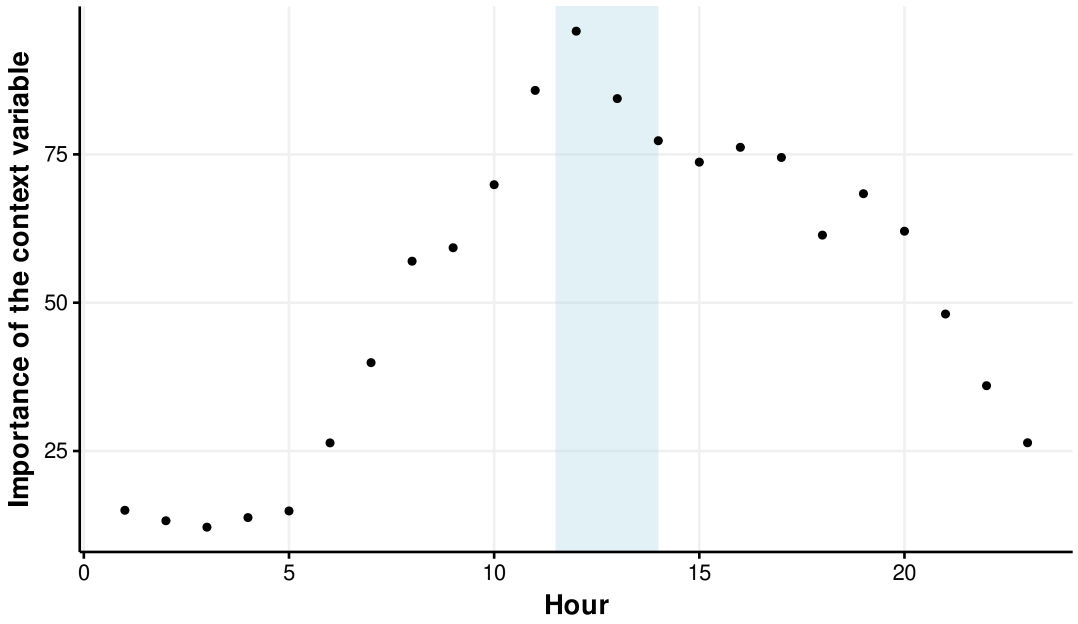

**Figure S5:** Importance of the FFO context variable $\phi(\mathbf{c}_{it})$ for models built with actions at different hours of the data. Variable importance is given by the absolute value of the $t$-statistic of the variable in the regression model (2) by the hour.

Each census tract can be described then by the set of frequencies of each topic within them. Most of the census tracts cannot be described uniquely by a single topic but by a weighted composition of them. As expected, we find that topics like “University” or “Airport” or “Seaside” are less frequent in census tracts than others like “Garage”, “Shopping Center” or “Neighborhood”, see figure S8.

In summary, our LDA method allowed us to characterize each census tract by the patterns or groups of different POIs we could find there. It is interesting to see that the POIs in almost twenty thousand census tracts can be described by only 20 different groups of POIs that co-occur frequently in our urban areas. Most of those groups/topics are related to commercial activities, but some of them describe working places, recreational or health areas too.

### 9 Effect of interventions on other health groups

Here we investigate the possibility that our different interventions target only specific health groups. To this end, we use the PLACES Local data for Better Health dataset from the CDC [5] that contains model-based census tract-level estimates for different health risk behaviors and outcomes. Using those estimates, we classify each individual into different groups of health outcomes depending on which quantile their census tract is with respect to that outcome. Figure S9 shows the total increase in the number of non-FFO visits for all interventions by health outcome group: Obesity, Diabetes, Sleeping less than 7 hours, Levels of Physical Inactivity, and High Cholesterol. As we can see, the environmental-behavioral intervention always increases the number of non-FFO visits across different groups with respect to the rest of the interventions. For example, compared with other interventions,

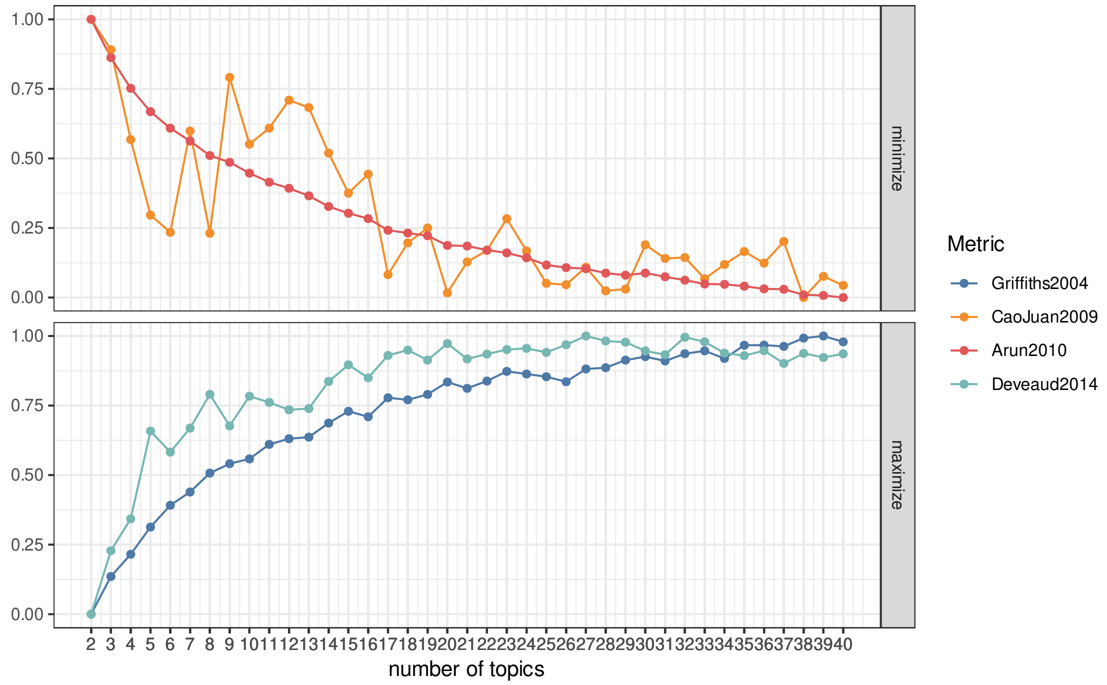

**Figure S6:** Variation of different metrics to estimate the optimal number of topics in the different census tracts. Metrics were obtained using the `ldatuning` package [18]. As we can see, several metrics show a minimum/maximum around $k = 20$.

we get 2.2x-2.5x more non-FFO visits on our environmental-behavioral intervention for groups coming from areas of high prevalence of diabetes and obesity. Note, however, that some other interventions have the same or even less effect on high-risk groups. For example, the "Food Swamp" and "Food Hotspots" interventions yield similar (low) effects in the high obesity, and diabetes groups but very different in low obesity, and diabetes groups. These results show that, although some other interventions might have a heterogeneous impact across different health risk groups, our "Environmental-Behavioral" intervention always affects significantly those with high health risks and significantly more than all other interventions.

### 10 Robustness checks

#### 10.1 Different definitions of POI visit

Visits to POIs are determined by looking at the closest POI to any stay up to a radius of $d^{max} = 200$ meters. Although we set $d^{max} = 200$m, we find that the median distance of stays attributed to POI is 24.24 m (Interquantil Range, IQR, 12.21 - 49.17), and the median distance of a stay attributed to FO is 18.82 m (IQR, 9.74 - 30.335m) and the median distance of a stay attributed to FFO is 17.70m (IQR, 9.257 - 31.88m). All of them much smaller than $d^{max} = 200$m. In any case, we have tested the robustness of our results to this attribution of stays to POI by exploring other values of $d^{max}$ from 20m to 200m. Again, the actual median distance to FO for those other values is much smaller than $d^{max}$: median of 10.14m (IQR, 6.15 - 14.55m) for $d^{max} = 20$m, 15.42m (IQR, 8.48 - 25.57m) for

$d^{max} = 50$m, and 17.61m (IQR, 9.31 - 31.89m) for $d^{max} = 100$m. As we can see in Figure S10, we find very slightly fluctuations with respect to $d^{max}$ of our results on the effect of the context on Fast Food visits. These results indicate that our main findings are largely independent of and robust to the details of our approach for attributing stays to POI.

##### 10.2 Different definitions of the context

We define the context $\mathbf{c}_{it}$ before lunch time as the last stay before 11:30. We have also tested the sensitivity of our result to other definitions of the context before lunch. For example, we tested taking $\mathbf{c}_{it}$ as the longest stay before lunchtime, or the last one with a duration longer than 1 hour. The results for the logistic regression are presented in Figure S10 where we can see that different definition of $\mathbf{c}_{it}$ yield very similar results on the effect of the context in fast food visits. These results indicate that our main findings are largely independent of and robust to the details of the definition of the context.

##### 10.3 Different characterization of the context

To characterize the mobile food environment through the variable $\phi(\mathbf{x})$ we have taken the ratio of FFO to FO in a radius of 1km around place $\mathbf{x}$. However, in highly dense areas with many FO, a radius of 1km can include many FO, and it will be less likely that people are using the whole information about that large area when making their decision. Thus, we have tested the sensitivity of our results to other definitions of $\phi(\mathbf{x})$ which account for the different densities of FO in different areas. In particular, we have tested taking $\phi(\mathbf{x})$ as the ratio of FFO to FO of the closest 25 FO to $\mathbf{x}$, a definition that takes into account the density of FO around a place. As we can see in Figure S10 our results are very similar for both definitions. These results indicate that our main findings are largely independent of and robust to the details of characterization of the context.

##### 10.4 Different sets of FO and FFO

Since there is little consensus about what is the definition of FFO, we test the robustness of our results to different definitions of food and fast food venues. In particular, we use three different definitions:

- WF (Wikipedia and Foursquare) definition, the one used in the main paper and described in Section 3.
- MF (Manual and Foursquare) definition, the one used in [13] for the Los Angeles metropolitan area, see Section 3.
- WF+ (Wikipedia and Foursquare extended), which is similar to the WF definition but also adds in all independent restaurants that have the words "pizza" or "burger" in their names, as was done in [7].

As we can see in Figure S11, our results for the effect of the mobile food environment on the individual decision to visit FFO are very similar across the different definitions. We note, however, that the effect seems to be larger for the MF definition, due to the restrictive definition made in [13] for an outlet to be considered as FFO. This makes the variable $\phi$ smaller in the different contexts and thus, the coefficient in the model gets larger.

#### 11 Interventions

We want to identify those areas to intervene by changing how individuals evaluate the options around them. For example, this can be done by changing the number of FFO around them or by ranking differently the restaurants in the area on recommendation platforms. Here we want to identify those areas where those changes could have more impact. Our model predicts that the probability of choosing FFO is given by

$$P(y_{it} = 1) = \frac{1}{1 + e^{\beta_0 - \delta_t - \alpha_i - \beta\phi(\mathbf{c}_{it})}}. \tag{4}$$

We want to investigate those places in which the change in that probability is larger after implementing an intervention I, that is, where the derivative with respect to I

$$\frac{\delta P(y_{it} = 1)}{\delta \mathrm{I}} = \beta \frac{e^{X_{it}}}{(1 + e^{X_{it}})^2} \frac{\delta \phi}{\delta \mathrm{I}} \tag{5}$$

is larger. Here $X_{it} = \beta_0 + \delta_t + \alpha_i + \beta\phi(\mathbf{c}_{it})$. If we focus on a specific area $\Omega$, then $\mathbf{c}_{it}$ is more or less the same within an area $\Omega$, then we get that the change in that area is

$$\Delta^{\mathrm{FFO}}(\Omega) = \sum_{\mathbf{c}_{it} \in \Omega} \beta \frac{e^{X_{it}}}{(1 + e^{X_{it}})^2} \frac{\delta \phi}{\delta \mathrm{I}} \tag{6}$$

This expression tells us that the change in probability of going to FFO in an area after changing the context around it depends on three things: i) the people that go to that area and their preferences (through $\alpha_i$), ii) also on the number of actions there, and of course, iii) the context around that area. For example, we can have an area in which many people go, but they have a large individual preference for non-FFO ($\alpha_i \ll 0$ ), and they are not influenced by the FFO around. In that case, the impact of the intervention I will be small. Or an area in which not many people go but they are very influenced by the FFO around ($\alpha_i \simeq 0$), so an intervention can change a large number of decisions.

##### Turn a Non-FF into a FF restaurant

Suppose we have an intervention where we turn just a FFO into a non-FFO in area $\Omega$. In that case $\phi(\mathbf{c}_{it})$ will change approximately by $-1/n_\Omega$, where $n_\Omega$ is the number of FO in an area $\Omega$. Therefore we get that $\delta\phi/\delta\mathrm{I} \simeq -1/n_\Omega$ and the total reduction in the number of FFO visits of that intervention in the area $\Omega$ is

$$\Delta^{\mathrm{FFO}}(\Omega) = \sum_{\mathbf{c}_{it} \in \Omega} \frac{\delta P(y_{it} = 1)}{\delta \mathrm{I}} = - \sum_{\mathbf{c}_{it} \in \Omega} \frac{\beta}{n_\Omega} \frac{e^{X_{it}}}{(1 + e^{X_{it}})^2} \tag{7}$$

#### 12 Libraries used

Analysis was conducted in R [20] using the following packages:

- Logistic and least square regressions with fixed effects were done using the `fixest` library [2].
- BSTS models were implemented using the `CausalImpact` package [4].
- Change of context was done using the `changepoint` [16]

- Packages `ggplot2` [27], and `tmap` [22] were used in the visualizations.
- Spatial analysis was done using the `sf` [19] package.
- LDA topic modeling was done using the `topicmodels` package [10].
- Approximate string matching for the classification of food outlets was done using the `stringdist` package [24].
- Access to the Census API was done using the `tidycensus` [26] package. Boundaries of the Census Block Groups were obtained from the Census API using the `tigris` [25] package.
- Regression tables were prepared using the `stargazer` package [12].

### References

[1] United States Census Bureau. Core-Based Statistical Areas. `https://www.census.gov/topics/housing/housing-patterns/about/core-based-statistical-areas.html`, 2000. Accessed: 22-06-2019.

[2] Laurent Bergé. Efficient estimation of maximum likelihood models with multiple fixed-effects: the R package FENmlm. *CREA Discussion Papers*, (13), 2018.

[3] David M Blei, Andrew Y Ng, and Michael I Jordan. Latent dirichlet allocation. *Journal of machine Learning research*, 3(Jan):993–1022, 2003.

[4] Kay H. Brodersen, Fabian Gallusser, Jim Koehler, Nicolas Remy, and Steven L. Scott. Inferring causal impact using bayesian structural time-series models. *Annals of Applied Statistics*, 9:247–274, 2015.

[5] Centers for Disease Control and Prevention. 500 Cities: local data for better health. `https://www.cdc.gov/places/about/500-cities-2016-2019/index.html`, 2017. Accessed: 08-01-2021.

[6] William Cohen, Pradeep Ravikumar, and Stephen Fienberg. A comparison of string metrics for matching names and records. In *Kdd workshop on data cleaning and object consolidation*, volume 3, pages 73–78, 2003.

[7] Janet Currie, Stefano DellaVigna, Enrico Moretti, and Vikram Pathania. The Effect of Fast Food Restaurants on Obesity and Weight Gain. *American Economic Journal: Economic Policy*, 2(3):32–63, 2010.

[8] Ashlesha Datar, Amy Mahler, and Nancy Nicosia. Association of exposure to communities with high obesity with body type norms and obesity risk among teenagers. *JAMA Network Open*, 3(3):e200846–e200846, 2020.

[9] Foursquare. Foursquare Venue Category Hierarchy. `https://developer.foursquare.com/docs/build-with-foursquare/categories/`, 2020. Accessed: 09-12-2020.

[10] Bettina Grün and Kurt Hornik. topicmodels: An R package for fitting topic models. *Journal of Statistical Software*, 40(13):1–30, 2011.

[11] Ramaswamy Hariharan and Kentaro Toyama. Project lachesis: parsing and modeling location histories. In *International Conference on Geographic Information Science*, pages 106–124. Springer, 2004.

[12] Marek Hlavac. *stargazer: Well-Formatted Regression and Summary Statistics Tables*. Central European Labour Studies Institute (CELSI), Bratislava, Slovakia, 2018. R package version 5.2.2.

[13] Abigail L. Horn, Brooke M. Bell, Bernardo Garcia Bulle Bueno, Mohsen Bahrami, Burcin Bozkaya, Yan Cui, John P. Wilson, Alex Pentland, Esteban Moro, and Kayla de la Haye. Investigating mobility-based fast food outlet visits as indicators of dietary intake and diet-related disease. *medRxiv*, 2021.

[14] Infogroup. Infogroup US Historical Business Data, 2016.

[15] Rebecca Killick and Idris A. Eckley. changepoint: An R Package for Changepoint Analysis. *Journal of Statistical Software*, 58(3):1–19, 2014.

[16] Rebecca Killick, Kaylea Haynes, and Idris A. Eckley. *changepoint: An R package for changepoint analysis*, 2022. R package version 2.2.3.

[17] Esteban Moro, Dan Calacci, Xiaowen Dong, and Alex Pentland. Mobility patterns are associated with experienced income segregation in large US cities. *Nature Communications*, 12(1):4633, 2021.

[18] Murzintcev Nikita. *ldatuning: Tuning of the Latent Dirichlet Allocation Models Parameters*, 2020. R package version 1.0.2.

[19] Edzer Pebesma. Simple Features for R: Standardized Support for Spatial Vector Data. *The R Journal*, 10(1):439–446, 2018.

[20] R Core Team. *R: A Language and Environment for Statistical Computing*. R Foundation for Statistical Computing, Vienna, Austria, 2020.

[21] Matthew Salganik. *Bit by bit: Social research in the digital age*. Princeton University Press, 2019.

[22] Martijn Tennekes. tmap: Thematic maps in R. *Journal of Statistical Software*, 84(6):1–39, 2018.

[23] United States Census Bureau. 2013-2017 American Community Survey 5-year Estimates. `https://www.census.gov/programs-surveys/acs`, 2019. Accessed: 2020-12-04.

[24] M.P.J. van der Loo. The stringdist package for approximate string matching. *The R Journal*, 6:111–122, 2014.

[25] Kyle Walker. *tigris: Load Census TIGER/Line Shapefiles*, 2020. R package version 1.0.

[26] Kyle Walker and Matt Herman. *tidycensus: Load US Census Boundary and Attribute Data as 'tidyverse' and 'sf'-Ready Data Frames*, 2020. R package version 0.10.2.

[27] Hadley Wickham. *ggplot2: Elegant Graphics for Data Analysis*. Springer-Verlag New York, 2016. R package version 3.3.2.

**Table S2:** Combinations of other parent category (not "Food") and category used to classify venues as food outlets.

| Parent Category | Category |
|---|---|
| Nightlife Spot | Sports Bar, Bar, Pub, Dive Bar, Speakeasy, Hookah Bar, Lounge, Seafood, American, Cocktail, Italian, Brewery, Beer Garden, Hotel Bar, Wine Bar, Mediterranean, Restaurant, Whisky Bar, Mexican, New American, Tiki Bar, Sake Bar, Liquor Store, Pizza, Gastropub, Wings, Sushi, Steakhouse, Coffee Shop, Korean, Wine Shop, Burgers, BBQ, Winery, Café, Beer Bar, French, Beach Bar, Sandwiches, Japanese, Asian, Convenience Store, Piano Bar, Beer Store, Bakery, Grocery Store, Desserts, Gourmet, Deli / Bodega, Juice Bar, Food Truck, Candy Store, Food & Drink, Cafeteria, Butcher, Snacks, Cheese Shop, Corporate Cafeteria, Supermarket, Street Food Gathering |
| Shop & Service | Grocery Store, Convenience Store, Liquor Store, Butcher, Gourmet, Food & Drink, Health Food Store, Warehouse Store, Discount Store, Deli / Bodega, Fish Market, Wine Shop, Candy Store, Beer Store, Cheese Shop, Supermarket, Sports Bar, Coffee Shop, Sandwiches, Farmer's Market, Snacks, Seafood, Fruit & Vegetable Store, Chocolate Shop, Café, American, Italian, Restaurant, Desserts, Gastropub, Bakery, Juice Bar, Herbs & Spices Store, Pizza, Winery, Dive Bar, Steakhouse, Food Truck, Burgers, Beer Garden, Lounge, Smoothie Shop, New American, Street Food Gathering, Organic Grocery, Wine Bar, Korean, Bar, Hookah Bar, Pub, Japanese, Mexican, Cocktail, Brewery, Sushi, Asian, Mediterranean, Speakeasy, Beer Bar, Tiki Bar, Cafeteria, BBQ, Whisky Bar, French, Hotel Bar, Sake Bar |
| Arts & Entertainment | Bar, Italian, Mexican, Piano Bar, Fish Market, Coffee Shop, Pub, American, New American, Restaurant, Gastropub, Sports Bar, Farmer's Market, Pizza, Lounge, Dive Bar, BBQ, Café, Steakhouse, Sushi, Wine Bar, Mediterranean, Desserts, Food Truck, Brewery, Beer Garden, Juice Bar, Cocktail, Speakeasy, Candy Store, Beach Bar, Burgers, Snacks, Bakery, Cafeteria, Japanese, Grocery Store, Food & Drink, Winery, Whisky Bar, Hotel Bar, French, Seafood, Asian, Wine Shop, Sake Bar, Hookah Bar, Beer Bar, Deli / Bodega, Sandwiches, Liquor Store, Cheese Shop, Convenience Store |
| Professional & Other Places | Winery, Liquor Store, Lounge, Coffee Shop, French, Italian, Corporate Cafeteria, Wine Shop, Hotel Bar, Chocolate Shop, American, Farmer's Market, Speakeasy, Bar, Fruit & Vegetable Store, Corporate Coffee Shop, Seafood, Café, Burgers, Cafeteria, Convenience Store, Grocery Store, Asian, Food & Drink, Deli / Bodega, Pub, Bakery, Restaurant, Candy Store, Mexican, Wine Bar, Whisky Bar, Sports Bar, Warehouse Store, Cocktail, Pizza, BBQ, Brewery, Mediterranean, Desserts, Hookah Bar, Beer Garden, Discount Store, Health Food Store, Snacks, Sandwiches, Food Truck, Japanese, Supermarket, Dive Bar, Steakhouse, New American, Gourmet, Juice Bar, Beer Bar, Cheese Shop, Beer Store |
| Travel & Transport | Lounge, Food Truck, Bar, Hotel Bar, Cocktail, Deli / Bodega, American, Pizza, Brewery, Restaurant, Cheese Shop, Pub, Bakery, Beer Garden, New American, Mediterranean, BBQ, Café, Liquor Store, Sports Bar, Sushi, Mexican, Convenience Store, Burgers, Wine Bar, Coffee Shop, Dive Bar, Beach Bar, Speakeasy, Italian, Seafood, Steakhouse, Wings, Sandwiches, Winery, Gastropub, French, Grocery Store, Korean |
| College & University | Cafeteria, Bar, Coffee Shop, Italian, Convenience Store, Café, Food Truck, American, Burgers, Restaurant, Pub, Speakeasy, Gourmet, Deli / Bodega, Sandwiches, Dive Bar, Cocktail, Snacks, Wine Bar, Hookah Bar, Lounge, [illegible] |

**Table S3:** Logistic regression results for the model in Equation [2] for different groups of FO visits and maximum distance between the context and the action, $d^{\max}$. Numbers in parenthesis correspond to standard errors of the coefficients. We also report the Squared Correlation and Pseudo $R^2$ results for the logistic regression.

| Dependent Variable: | | | $y_{it}$ | | |
|---|---|---|---|---|---|
| Model: | All | $d^{\max} = 1km$ | $d^{\max} = 2km$ | $d^{\max} = 5km$ | $d^{\max} = 10km$ |
| *Variables* | | | | | |
| $\phi(\mathbf{c}_{it})$ | 1.872*** | 9.689*** | 6.536*** | 3.794*** | 2.726*** |
| | (0.0335) | (0.1158) | (0.0820) | (0.0541) | (0.0428) |
| *Fixed-effects* | | | | | |
| Day | Yes | Yes | Yes | Yes | Yes |
| User | Yes | Yes | Yes | Yes | Yes |
| *Fit statistics* | | | | | |
| Observations | 1,483,379 | 385,994 | 545,260 | 855,973 | 1,112,032 |
| Squared Correlation | 0.18820 | 0.27075 | 0.24201 | 0.21350 | 0.20060 |
| Pseudo $R^2$ | 0.16054 | 0.22632 | 0.20270 | 0.18010 | 0.17008 |
| BIC | 4,438,285.7 | 1,245,221.4 | 1,782,324.6 | 2,735,133.6 | 3,457,071.7 |

*Clustered (day & user) standard-errors in parentheses*
*Signif. Codes: ***: 0.01, **: 0.05, *: 0.1*

**Table S4:** Results for the Ordinary Least Squares regression of mobile ($\phi_i^m$), home ($\phi_i^h$) average exposure to FFO environments, and the fraction of FFO visits ($\mu_i$) by individual, as a function of different socio-demographic traits of the census tract where individuals live.

| | *Dependent variable:* | | |
|---|---|---|---|
| | $\phi^m$ | $\phi^h$ | $\mu$ |
| Median Income | 0.002*** (0.0002) | −0.019*** (0.0004) | −0.0002 (0.0005) |
| Proportion of Black | 0.022*** (0.0004) | 0.048*** (0.001) | 0.037*** (0.001) |
| Proportion of high education | −0.016*** (0.001) | 0.025*** (0.001) | −0.027*** (0.001) |
| Proportion employed | 0.007*** (0.001) | 0.027*** (0.003) | 0.005 (0.003) |
| Proportion using public transportation | −0.093*** (0.001) | 0.019*** (0.001) | −0.106*** (0.001) |
| Proportion with long commutes | 0.050*** (0.001) | −0.073*** (0.001) | 0.068*** (0.001) |
| Proportion employed in low skill jobs | 0.040*** (0.001) | 0.008*** (0.001) | 0.043*** (0.002) |
| Constant | 0.098*** (0.002) | 0.262*** (0.004) | 0.165*** (0.005) |
| Fixed effect by urban area | YES | YES | YES |
| Observations | 1,001,733 | 1,001,733 | 1,001,733 |
| $R^2$ | 0.213 | 0.038 | 0.052 |
| Adjusted $R^2$ | 0.213 | 0.038 | 0.052 |
| Residual Std. Error (df = 1001715) | 0.063 | 0.128 | 0.155 |
| F Statistic (df = 17; 1001715) | 15,921.330*** | 2,342.803*** | 3,240.151*** |
| *Note:* | | | *p<0.1; **p<0.05; ***p<0.01 |

**Table S5:** Logistic regression results for the model in Equation [2] for different groups of users and food outings. The last column corresponds to the model in Equation [3] for the DMV visits. Numbers in parenthesis correspond to standard errors of the coefficients. We also report the Squared Correlation and Pseudo $R^2$ results for the logistic regression.

| Dependent Variable: | $y_{it}$ | | | | | | | | | |
|---|---|---|---|---|---|---|---|---|---|---|
| Food Outings group: | All | Weekdays | Weekends | Income Q1 | Income Q2 | Income Q3 | Income Q4 | Low FFO | Mid FFO | High FFO |
| *Variables* | | | | | | | | | | |
| $\phi(\mathbf{c}_{it})$ | $1.842^{***}$ | $1.765^{***}$ | $1.766^{***}$ | $1.851^{***}$ | $1.781^{***}$ | $1.884^{***}$ | $1.871^{***}$ | $2.999^{***}$ | $1.817^{***}$ | $1.765^{***}$ |
| | (0.0391) | (0.0491) | (0.1191) | (0.0630) | (0.0711) | (0.0733) | (0.0916) | (0.1891) | (0.0608) | (0.0506) |
| $\hat{\alpha}_i$ | | | | | | | | | | |
| (Intercept) | | | | | | | | | | |
| *Fixed-effects* | | | | | | | | | | |
| Day | Yes | Yes | Yes | Yes | Yes | Yes | Yes | Yes | Yes | Yes |
| User | Yes | Yes | Yes | Yes | Yes | Yes | Yes | Yes | Yes | Yes |
| *Fit statistics* | | | | | | | | | | |
| Observations | 1,072,640 | 781,753 | 108,813 | 332,518 | 280,331 | 254,590 | 205,201 | 105,888 | 534,335 | 436,173 |
| Squared Correlation | 0.19190 | 0.19555 | 0.13246 | 0.19573 | 0.19272 | 0.19211 | 0.18745 | 0.13105 | 0.12432 | 0.17329 |
| Pseudo $R^2$ | 0.16221 | 0.16411 | 0.10513 | 0.16526 | 0.16286 | 0.16252 | 0.15867 | 0.14224 | 0.11201 | 0.13679 |

*Signif. Codes: ***: 0.01, **: 0.05, *: 0.1*

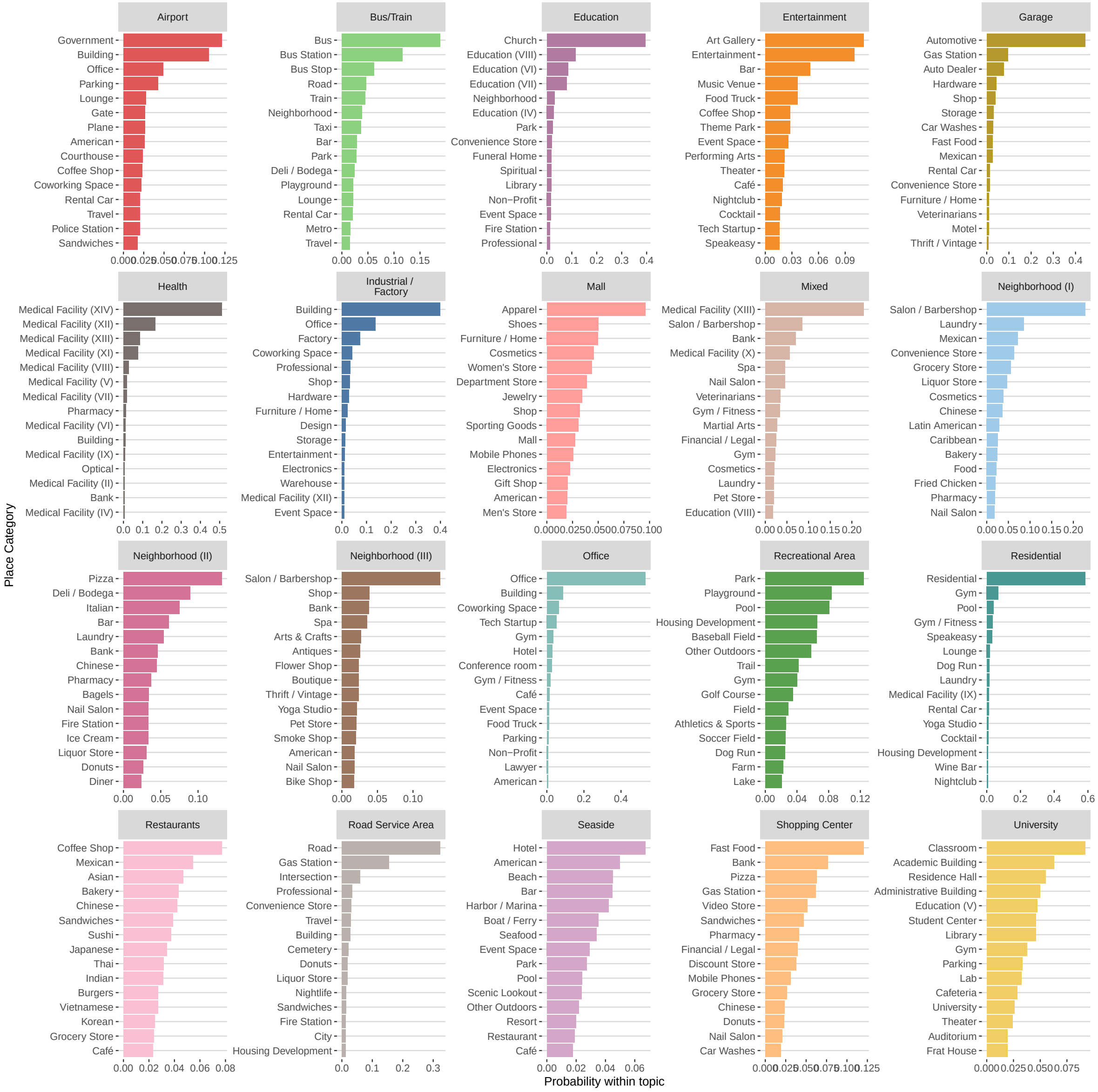

**Figure S7:** Composition of the different $k = 20$ POI topics found in our census tracts. We have given names to each of them for easy recognition. Bars represent the probability of finding each POI category in each of the topics. Note: no mobility data or visits were used in this analysis, only the spatial distribution of POIs from the Foursquare API.

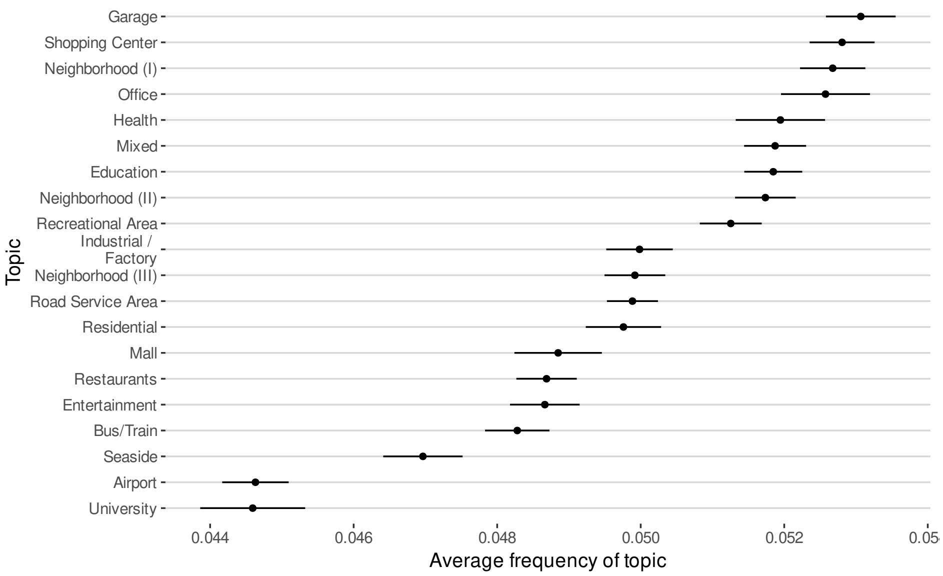

**Figure S8:** Average (and standard error of the mean) frequency of each topic across all census tracts in our dataset.

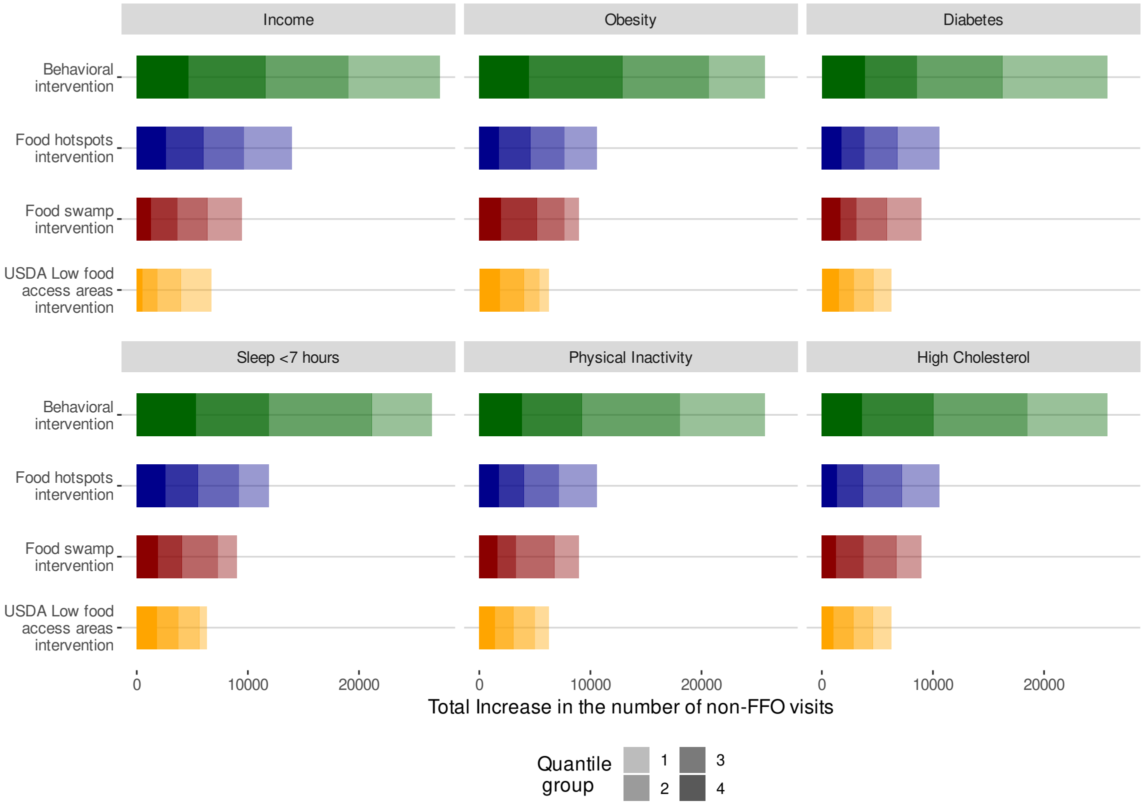

**Figure S9:** Effect of the intervention strategies on the different health outcomes groups. Each panel shows the total increase in the number of non-FFO visits by the changed actions of users belonging to different health outcomes groups (shades). We classify each individual into different groups of health outcomes depending on which quantile their census tract is with respect to that outcome (darker shading for higher quantiles of each health outcome group). Health outcomes by census tract are obtained from the PLACES Local data for Better Health dataset from the CDC [5].

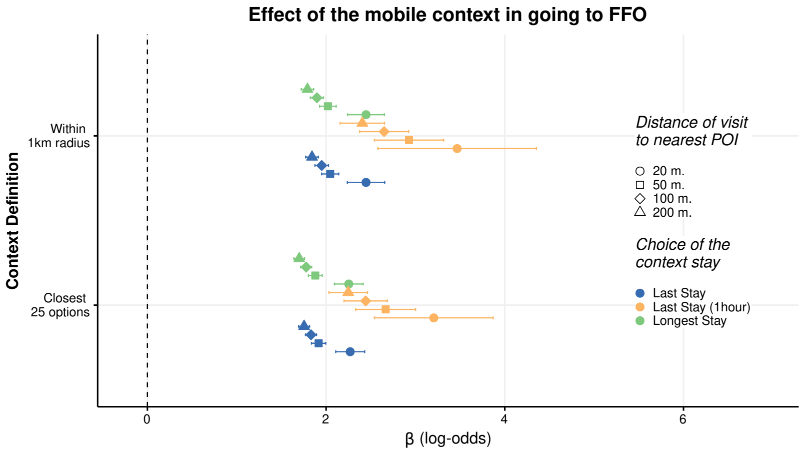

**Figure S10:** Effect of the mobile phone context in choosing a FFO when going to a food place at lunchtime for different definitions of FFO for different attributions of the stays to the POI and the choice of the stay for the context before lunch. Values show the coefficient $\beta$ of the logistic regression (log-odds) in Eq. [2] and bars indicate its standard deviation. The result in the main paper corresponds to 200m, within a 1km radius and last stay.

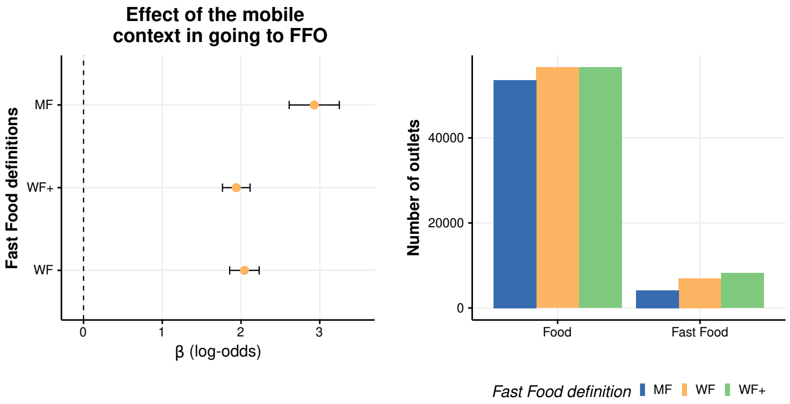

**Figure S11:** Left: Effect of the mobile phone context in choosing a FFO when going to a food place at lunchtime for different definitions of FFO. The analysis is only made for the Los Angeles metropolitan area since the manual definition in [13] is only available there. Values show the coefficient of the logistic regression (log-odds) in Eq. [2] and bars indicate its standard deviation. Right: Number of FO and FFO in the different definitions of fast food outlets.
